# Association Between Hydrogen-Rich Water Consumption and Lower Extremity Function in Older Adults Participating in Community Salons: A Prospective Observational Study

**DOI:** 10.64898/2026.01.07.26343642

**Authors:** Yuusuke Harada, Michiko Miyakawa

## Abstract

**Background:** Falls among older adults are a leading cause of fractures, loss of independence, and need for long-term care. Community salons in Japan promote social participation and health activities among older adults. Hydrogen-rich water is widely used as a health product, but evidence in community settings remains limited.

**Methods:** We conducted a prospective observational study among 48 community-dwelling older adults attending community salons in Hiroshima City, Japan. Hydrogen-rich water was offered by the salon operators as part of routine activities; the research team did not assign participants to consume it. Participants were categorized at baseline according to their usual hydrogen-rich water consumption at the salons (consumers vs non-consumers) and followed for six months. The primary outcome was the 30-second chair stand test (CS-30). Secondary outcomes included the Timed Up and Go test (TUG), usual gait speed, one-leg stance time, and grip strength. Within-group changes and between-group differences in change scores were compared.

**Results:** All 48 participants completed follow-up, and no serious adverse events were reported during the study period. The consumers group showed a greater improvement in CS-30 over six months (baseline 12.96 (SD 3.21) to follow-up 14.52 (SD 3.59); change 1.57 (SD 2.41)) compared with the non-consumers group (12.52 (SD 3.00) to 12.22 (SD 3.54); change −0.30 (SD 1.55)), with a significant between-group difference in change scores (p=0.003). The consumers group also showed a greater increase in usual gait speed (0.91 (SD 0.24) to 0.98 (SD 0.26); change 0.07 (SD 0.08)) than the non-consumers group (0.94 (SD 0.24) to 0.97 (SD 0.22); change 0.03 (SD 0.05); p=0.008). No significant between-group differences were observed for TUG (p=0.57), one-leg stance time (p=0.13), or grip strength (p=0.10).

**Conclusion:** In community-dwelling older adults participating in community salons, routine hydrogen-rich water consumption was associated with improved lower extremity function as measured by CS-30 and gait speed. Because exposure was not randomized, residual confounding cannot be excluded, and causal inference is limited. Larger studies with stronger designs are warranted.

## Introduction

With population aging, falls in older adults are a major contributor to fractures, loss of independence, and the need for long-term care [1]. Exercise-based fall prevention programs are effective in reducing fall risk among community-dwelling older adults [2]. Fall risk is multifactorial, but declines in lower-extremity strength and gait ability are key modifiable factors [3–5]. In Japan, community “salons” (also called “places to gather”) have been established across regions as resident-led venues where older adults engage in group exercise and social activities to prevent frailty and promote participation [6]. However, it remains unclear to what extent such low-to-moderate intensity community programs can sustainably improve physical function, and whether combining exercise with non-exercise components could enhance the effectiveness of pragmatic interventions.

Hydrogen-rich water containing molecular hydrogen (H2) has attracted interest for its selective antioxidant and anti-inflammatory properties. A recent systematic review suggests that evidence for performance and fatigue-related outcomes is mixed across populations and protocols [7].

Studies in younger populations and athletes suggest that hydrogen-rich water may reduce exercise-induced fatigue and markers of oxidative stress [8], and in individuals with lifestyle-related conditions, improvements in oxidative stress and metabolic markers have been reported [9]. In older adults, a six-month randomized controlled pilot trial reported that hydrogen-rich water intake may improve aging-related biomarkers such as telomere length, inflammatory markers, and lower-extremity muscle strength [10]. Nevertheless, evidence focusing on functional outcomes relevant to falls (e.g., chair stand performance, mobility tests, and gait speed) in community-based older populations is limited.

Therefore, we conducted a prospective observational study in older adults attending community salons in Japan. All participants engaged in the same routine community salon activities, including regular group exercise classes and encouragement of self-directed outdoor walking.

We compared changes in fall-related functional indicators between participants who routinely consumed hydrogen-rich water available at the salons and those who did not. We hypothesized that routine consumption of hydrogen-rich water would be associated with greater improvements in lower-extremity function. The primary outcome was the 30-second chair stand test (CS-30), and secondary outcomes were TUG, usual gait speed, one-leg stance time, and grip strength.

## Methods

### Study design and setting

This was a prospective observational study conducted in community salons in Hiroshima City, Japan. Hydrogen-rich water was offered by the salon operators as part of routine activities; the research team did not assign, prescribe, or provide hydrogen-rich water for research purposes.

### Participants

Participants were 48 Japanese older adults who regularly attended community salons. Individuals who were able to participate in the assessments and who provided informed consent were enrolled. The overall mean age was 75.9±4.9 years. Exclusion criteria were: (1) certification as requiring support or long-term care; (2) severe cognitive impairment that precluded valid assessment; (3) conditions that made the assessments unsafe; and (4) other reasons deemed inappropriate by the principal investigator.

### Exposure assessment

Hydrogen-rich water consumption status was assessed at baseline based on usual consumption at the community salons. As part of the routine salon program, hydrogen-rich water packaged in aluminum pouches was distributed on salon days. Participants who chose to consume it took home a supply corresponding to the number of days until the next salon session. Participants were categorized as consumers if they routinely consumed the hydrogen-rich water and as non-consumers if they did not. The hydrogen concentration of the product used in the program was reported to be approximately 1.05 ppm, and intake volume was typically up to 600 mL/day among consumers.

### Ethics

This study was approved by the Chiba University Ethics Committee (Approval No. M846). The data used were anonymized, and study information was publicly disclosed to provide an opt-out opportunity.

### Outcomes and assessments

The primary outcome was CS-30, an indicator of lower-extremity strength and fall risk. Participants were instructed to stand up from and sit down on a chair as many times as possible in 30 seconds; the completed repetitions were recorded. Secondary outcomes were grip strength, TUG, usual gait speed, and one-leg stance time.

### Statistical analysis

Continuous variables are presented as mean±SD and categorical variables as n (%). Within-group changes from baseline to six months were assessed, and between-group differences in change scores were compared using independent-samples t-tests. Statistical significance was set at p<0.05. The sample size was determined by feasibility rather than an a priori power calculation.

## Results

### Participants

A total of 48 community-dwelling older adults were enrolled and categorized at baseline (hydrogen-rich water consumers: 24; non-consumers: 24). Baseline characteristics were similar between groups (Table 1). All 48 participants completed six-month follow-up. No serious adverse events were reported during follow-up.

**Table 1.** Baseline Characteristics of Participants by Hydrogen-Rich Water Consumption Status (N=48)

| Variable | Hydrogen-rich water consumers (n = 24) | Non-consumers (n = 24) | p-value |
| --- | --- | --- | --- |
| Age, years | 76.3 ± 4.9 | 75.4 ± 5.0 | 0.52 |
| Female, n (%) | 17 (70.8%) | 14 (58.3%) | 0.55 |
| History of falls in past year, n (%) | 11 (45.8%) | 11 (45.8%) | 1.00 |
| Any walking aid (cane or walker), n (%) | 8 (33.3%) | 7 (29.2%) | 1.00 |
| CS-30, repetitions | 13.0 ± 3.2 | 12.5 ± 2.9 | 0.51 |
| TUG, s | 10.9 ± 2.5 | 11.0 ± 2.5 | 0.95 |
| Usual gait speed, m/s | 0.92 ± 0.24 | 0.91 ± 0.22 | 0.85 |
| One-leg stance, s | 15.7 ± 13.9 | 14.7 ± 13.6 | 0.81 |
| Grip strength, kg | 20.7 ± 7.1 | 22.1 ± 7.4 | 0.50 |

Values are mean ± SD or n (%).
P-values are from independent-samples t-tests for continuous variables and Fisher's exact tests for categorical variables.

### Physical function outcomes

Changes in physical function over six months are shown in Table 2. The consumers group demonstrated a greater improvement in CS-30 compared with the non-consumers group (p=0.003). The consumers group also showed a greater increase in usual gait speed (p=0.008). Between-group differences were not significant for TUG (p=0.57), one-leg stance time (p=0.13), or grip strength (p=0.10).

**Table 2.** Changes in Physical Function Over Six Months by Hydrogen-Rich Water Consumption Status.

| Outcome | Group | n | Baseline mean ± SD | Six-month mean ± SD | Change (Six months - Baseline) mean ± SD | p-value |
| --- | --- | --- | --- | --- | --- | --- |
| CS-30, repetitions | Consumers | 23 | 12.96 ± 3.21 | 14.52 ± 3.59 | 1.57 ± 2.41 | 0.003 |
| | Non-consumers | 23 | 12.52 ± 3.00 | 12.22 ± 3.54 | -0.30 ± 1.55 | |
| TUG, s | Consumers | 23 | 10.93 ± 2.60 | 10.72 ± 3.97 | -0.22 ± 3.28 | 0.57 |
| | Non-consumers | 23 | 11.00 ± 2.59 | 11.18 ± 2.67 | 0.18 ± 0.44 | |
| Usual gait speed, m/s | Consumers | 23 | 0.91 ± 0.24 | 0.98 ± 0.26 | 0.07 ± 0.08 | 0.008 |
| | Non-consumers | 23 | 0.91 ± 0.22 | 0.91 ± 0.22 | -0.00 ± 0.08 | |
| One-leg stance, s | Consumers | 22 | 15.95 ± 14.16 | 17.00 ± 15.64 | 1.05 ± 4.63 | 0.13 |
| | Non-consumers | 23 | 13.30 ± 11.95 | 12.48 ± 12.39 | -0.83 ± 3.47 | |
| Grip strength, kg | Consumers | 23 | 20.87 ± 7.21 | 21.04 ± 7.71 | 0.17 ± 1.61 | 0.10 |
| | Non-consumers | 23 | 21.52 ± 7.01 | 20.91 ± 7.21 | -0.61 ± 1.59 | |

P-values are from independent-samples t-tests comparing change scores between groups.
Note: For some outcomes, the analytic sample size was 22-23 due to occasional inability to complete specific assessments.

## Discussion

### Principal findings

In this prospective observational study of older adults attending community salons, routine hydrogen-rich water consumption was associated with greater improvements in CS-30 and usual gait speed over six months compared with non-consumption. No significant between-group differences were observed for TUG, one-leg stance time, or grip strength.

### Interpretation

CS-30 reflects lower-extremity strength and functional mobility, which are important determinants of fall risk. The observed associations may reflect differences in lifestyle, health behaviors, or baseline motivation between consumers and non-consumers in addition to any potential biological effects of hydrogen-rich water. Hydrogen-rich water has been proposed to exert antioxidant and anti-inflammatory effects, but evidence in older adults remains limited. Randomized studies in older adults have reported improvements in selected physical function outcomes with hydrogen-containing water, although findings vary by protocol and outcome [10,11]. Experimental work suggests potential mechanisms via oxidative stress modulation and metabolic pathways, but translation to pragmatic community settings remains uncertain [12].

Evidence in physically active adults also suggests possible effects on fatigue-related outcomes, supporting ongoing investigation across populations [13]. Proposed mechanisms include signaling pathways such as ghrelin-related regulation reported in prior studies [14,15].

### Strengths and limitations

A strength of this study is its pragmatic community setting, with complete follow-up and repeated objective assessments of physical function.

Several limitations should be considered. Participants were community salon attendees who were able to attend group activities, suggesting relatively preserved mobility; therefore, generalizability to frailer, institutionalized, or homebound older adults is limited. We did not comprehensively assess dietary intake or broader health-related factors (e.g., nutrition, supplement use, comorbidities, medications, and physical activity), and residual confounding cannot be excluded. Exposure classification relied on routine intake within the program and may be subject to misclassification; intake outside the salon and the hydrogen concentration at the time of consumption were not directly measured. Moreover, the dissolved hydrogen concentration reported in a prior trial [10] appears higher than would be expected under atmospheric pressure, raising concerns about differences in measurement conditions or labeling and limiting comparability across studies. Finally, the sample size was modest, which may have reduced statistical power to detect small-to-moderate associations. As a non-randomized observational study, causal inference is limited.

### Implications

Community salons are a key platform for fall prevention and healthy aging. If future studies confirm benefits, hydrogen-rich water consumption could be considered as a supportive component of community-based health promotion programs.

## Conclusions

In a prospective observational study in community-dwelling older adults attending community salons, routine hydrogen-rich water consumption was associated with improved lower extremity function as assessed by CS-30 and usual gait speed over six months. Because participants were not randomly assigned to exposure, unmeasured confounding may have influenced the findings. Larger studies with stronger designs are warranted.

## Data Availability

All data produced in the present study are available upon reasonable request to the authors

